# Inheritance of a common androgen synthesis variant allele is associated with female COVID susceptibility in UK Biobank

**DOI:** 10.1101/2020.08.27.20183004

**Authors:** Jeffrey M. McManus, Navin Sabharwal, Peter Bazeley, Nima Sharifi

## Abstract

**Context:** A sex discordance in COVID exists, with males disproportionately affected. Although sex steroids may play a role in this discordance, no definitive genetic data exist to support androgen-mediated immune suppression for viral susceptibility, nor for adrenally produced androgens.

**Objective:** The common adrenal-permissive missense-encoding variant *HSD3B1*(1245C) that enables androgen synthesis from adrenal precursors and that has been linked to suppression of inflammation in severe asthma was investigated in COVID susceptibility and outcomes reported in the UK Biobank.

**Methods:** The UK Biobank is a long-term study with detailed medical information and health outcomes for over 500,000 genotyped individuals. We obtained COVID test results, inpatient hospital records, and death records and tested for associations between COVID susceptibility or outcomes and *HSD3B1*(1245A/C) genotype. The outcomes were identification as a COVID case among all subjects, COVID positivity among COVID-tested subjects, and mortality among subjects identified as COVID cases.

**Results:** Adrenal-permissive *HSD3B1*(1245C) genotype was associated with identification as a COVID case (odds ratio 1.11 per C allele, p = 0.00054) and COVID test positivity (OR 1.10, p = 0.0036) in older (≥ 70 years of age) women. In women identified as COVID cases, there was a positive linear relationship between age and 1245C allele frequency (p < 0.0001). No associations were found between genotype and mortality.

**Conclusion:** Our study suggests that a common androgen synthesis variant regulates immune susceptibility to COVID infection in women, with increasingly strong effects as women age.

## Introduction

The COVID-19 pandemic caused by the SARS-CoV-2 virus emerged in late 2019 and in the early months of 2020 quickly became a global health emergency. Recognition that men face higher risks of hospitalization and death from COVID than women^1-3^ has spurred research into whether sex hormone signaling plays a role in COVID susceptibility and/or outcomes. Although much of this research has focused on whether androgens regulate viral receptor protein ACE2 and co-receptor TMPRSS2^4-8^, it is also known that sex hormone signaling can modulate immune responses^9-11^. Generally speaking, a sex discordance in inflammatory and infectious disease processes^12,13^, including COVID, is recognized. However, there is great uncertainty in attributing the difference in outcomes to androgens because there are many other physiologic differences that exist between males and females.

The enzyme 3β-hydroxysteroid dehydrogenase isotype 1 (3βHSD1) catalyzes a critical and rate-limiting step in the pathway to production in peripheral tissues of the potent androgens testosterone and dihydrotestosterone (DHT) from adrenally produced precursors dehydroepiandrosterone (DHEA) and its sulfated form DHEA-S^14^. The adrenal-permissive form of the gene for 3βHSD1, *HSD3B1*(1245C), was originally linked to faster prostate cancer progression in the face of androgen deprivation therapy (i.e., medical castration) because of adrenally derived androgen synthesis^15-20^. More recently, *HSD3B1* genotype has been shown to also affect the response to oral glucocorticoid treatment in patients with severe asthma^21^, an inflammatory condition of the lungs often associated with T cell activation^22^. The 1245C allele encodes a form of the enzyme that is resistant to ubiquitination and degradation, leading to increased production of potent androgens from adrenal precursors, and can therefore be described as adrenal androgen permissive, or “adrenal-permissive”^23^ for simplicity. By contrast, the 1245A, or adrenal-restrictive allele, encodes a more rapidly degraded form of the enzyme, leading to less production of potent androgens^24^.

A side effect of glucocorticoid treatment is suppression of circulating DHEA and DHEA-S levels^21^. Active androgens (i.e., potent agonists of the androgen receptor) testosterone and DHT are thought to downregulate immune/inflammatory responses^25^. Experimental evidence for androgen-induced immunosuppression includes, for example, that castration of mice reduces B cell lymphopoiesis and this is reversed by androgen replacement^26^ and that androgen ablation allows greater T cell response to prostate tumors in a mouse immunotherapy model^27^. In asthma patients, higher androgen receptor expression in the bronchial epithelium has been associated with lower disease severity^28^, and inherited androgen receptor deficiency has been associated with greater risk of asthma^29^. Thus, in the asthma context, even in an environment of a decreased adrenal precursor pool brought on by glucocorticoid treatment, the adrenal-permissive 1245C allele still enables sufficient downstream androgen production to reduce airway inflammation in those with severe asthma, whereas patients with the adrenal-restrictive 1245A allele experience less reduction in airway inflammation^21^ (**Fig. 1**).

**Figure 1.**
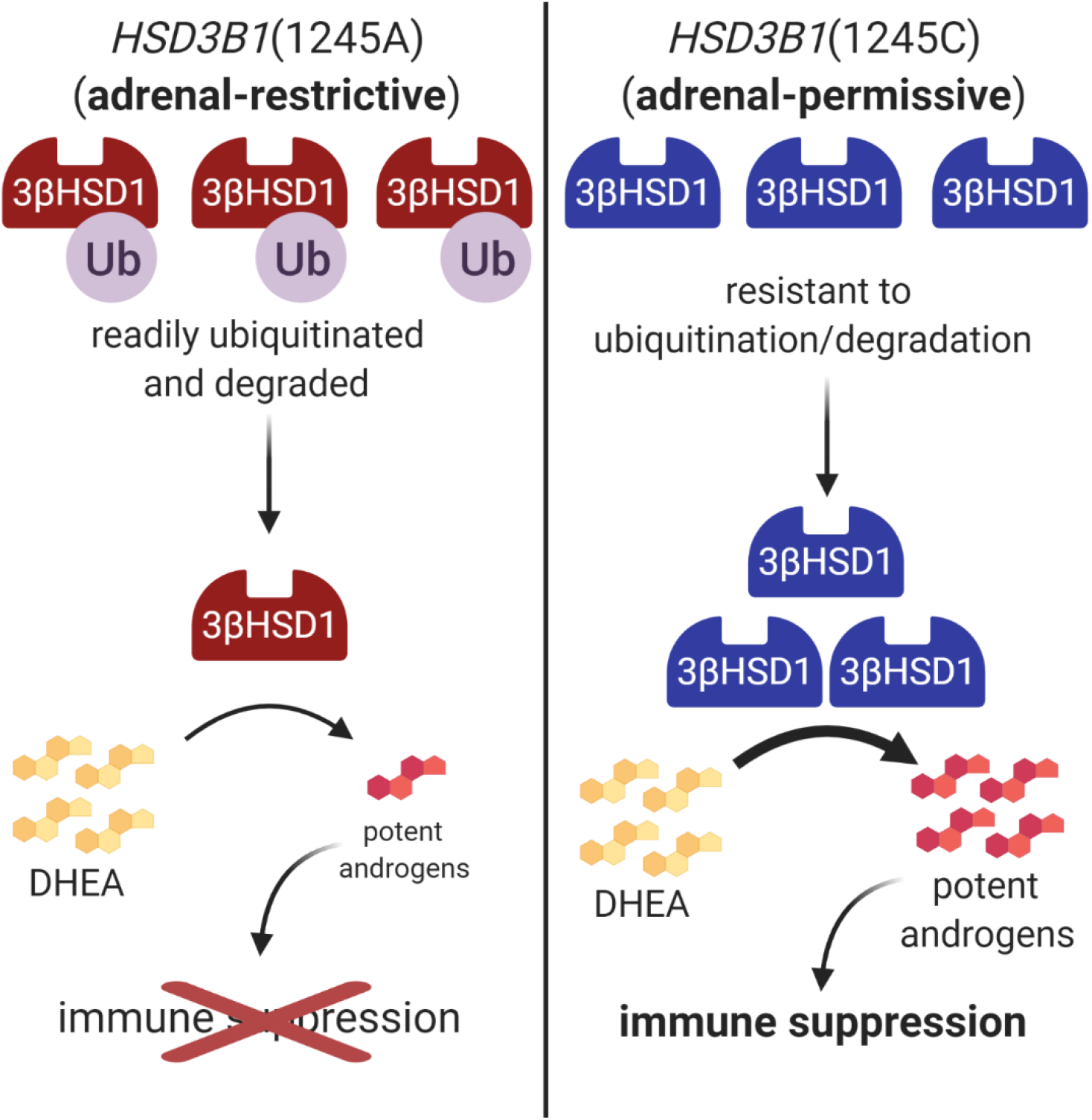
Hypothesized mechanism: adrenal-permissive *HSD3B1* genotype leads to greater androgen synthesis and immune suppression. Depending on genotype, *HSD3B1* codes for an enzyme 3βHSD1 that is either readily ubiquitinated and degraded (adrenal-restrictive form, left) or is resistant to ubiquitination and degradation (adrenal-permissive form, right) and as a result accumulates at higher levels in cells. The higher levels of the adrenal-permissive enzyme could enable greater conversion of DHEA to downstream potent androgens at sufficient levels to suppress immune responses, whereas these levels would not be reached with the adrenal-restrictive form. Created with BioRender.com.

The evidence from cohorts of severe asthmatics that *HSD3B1* genotype could affect function of the immune system led us to ask whether there are links between *HSD3B1* genotype and COVID susceptibility and/or outcomes. Additionally, in both prostate cancer and asthma, the effect of the adrenal-permissive allele becomes evident specifically in contexts of decreased circulating androgen levels due to androgen deprivation therapy in prostate cancer and glucocorticoid treatment in asthma. Circulating androgen levels decline with age^30-32^, so as COVID most severely impacts older patients^2,33^, this provided further impetus for determining whether a link exists between *HSD3B1* and COVID.

## Materials and Methods

Analysis was performed on subjects from the UK Biobank, a long-term study tracking medical information of over 500,000 genotyped individuals. Genotyping was performed using the UK BiLEVE Axiom Array or UK Biobank Axiom Array by Applied Biosystems. Additional SNPs were imputed with the IMPUTE4 program using the Haplotype Reference Consortium, UK10K, and 1000 Genomes phase 3 panels as reference panels. Rs1047303 (i.e., *HSD3B1*(1245A/C)) was imputed with high accuracy (information score > 0.99).

Population structure and ancestry were assessed via principal component analysis on SNP data. Final sample sizes for different cohorts as defined by the UK Biobank after quality control, among subjects with *HSD3B1* genotyping and who were alive as of the beginning of 2020, were 362,441 Caucasian, 6,975 African, 6,898 South Asian, 1,372 Chinese, and 82,218 unclassified. Analyses were performed both on this full set of subjects (n = 459,904) and on the UK Biobank-defined Caucasian cohort (n = 362,441), the latter to verify that results were not being confounded by race or ethnicity.

COVID test results, inpatient hospital records, and death records from the UK Biobank were downloaded. As of April 2021, COVID test results up to March 15, 2021, hospital admissions up to February 2, 2021, and death records up to March 23, 2021 were available and were included in analyses. Subjects with COVID test results were identified and classified as having tested positive or having solely negative test results. Subjects with ICD10 codes U07.1 (i.e., laboratory-confirmed COVID) or U07.2 (i.e., clinically diagnosed COVID without laboratory confirmation) from inpatient hospital episodes were also identified and subjects with COVID ICD10 codes despite having no or solely negative COVID test results were included as additional COVID cases. For analysis of mortality among subjects who had COVID, death records were cross referenced against COVID test results and ICD10 codes and subjects who died at any time after having tested positive for COVID and/or having been diagnosed with COVID as a hospital inpatient were identified; i.e., all-cause mortality for subjects who had COVID was assessed. Data on which UK Biobank subjects received glucocorticoid treatment while hospitalized with COVID were not available.

Blood samples were collected from UK Biobank subjects upon enrollment to the study in 2006-2010 and key biochemistry markers were measured, including testosterone and estradiol, which were measured using a Beckman Coulter DXI 800 with analysis methodology chemiluminescent immunoassay – competitive binding. The bottom of the reportable range was 0.35 nM for testosterone and 175 pM for estradiol.

Analyses were performed using R under RStudio. This study was approved by the NHS National Research Ethics Committee (REC ref 16/NW/0274) through the generic ethical approval for UK Biobank studies (approved UK Biobank application #44578).

## Results

To examine whether COVID susceptibility and *HSD3B1* genotype are associated, we performed regression analyses between identification as a COVID case and genotype in all genotyped subjects of the UK Biobank who were alive at the beginning of 2020. The UK Biobank is a long-term study containing genotyping results and detailed medical information from approximately 500,000 UK residents who were between the ages of 40 and 69 years when recruited to the study in 2006-2010^34^. Current ages (as of September 2020, the midpoint of the COVID results) range from 48 to 86 with a median age of 69. **Fig. S1**^35^ shows a histogram of the ages of UK Biobank subjects included in the analysis; over 99.5% of subjects had ages within the range 51 – 82. As of the beginning of April 2021, COVID test results had been reported for 69,349 genotyped UK Biobank subjects, 15,812 having tested positive. An additional 513 subjects who did not have positive test results were identified as COVID cases by ICD10 codes from inpatient hospital episodes, bringing the total number of cases to 16,325.

**Table 1** shows the characteristics of UK Biobank subjects who were identified as COVID cases and of the rest of the subject population, who served as controls. Frequencies of *HSD3B1*(1245C) vary by race, ranging in the UK Biobank from 0.074 and 0.085 in the Chinese and African cohorts, respectively, to 0.193 in the South Asian cohort and 0.323 in the Caucasian cohort, which contains over 75% of UK Biobank subjects. Our regression analyses were adjusted for the first ten genetic principal components, but to further ensure that ancestry was not a confounding factor in the results, we also performed the same analyses but restricted to the Caucasian cohort as defined by the UK Biobank. As shown in Table 1, in the UK Biobank population, subjects identified as COVID cases were more likely to be male, younger on average, more likely to be of non-White ethnicity, and had higher BMI on average than controls.

**Table 1.** Characteristics of UK Biobank subjects identified as COVID cases and subjects who ved as controls.

|  | All UK Biobank subjects who were alive in 2020 |  | UK Biobank's Caucasian cohort subjects who were alive in 2020 |  |
| --- | --- | --- | --- | --- |
|  | COVID cases<br>(n = 16,325) | Controls<br>(n = 443,579) | COVID cases<br>(n = 12,031) | Controls<br>(n = 350,410) |
| **Sex** |  |  |  |  |
| **Male** | 7,763 (47.6%) * | 198,890 (44.8%) | 5,786 (48.1%) * | 158,769 (45.3%) |
| **Female** | 8,562 (52.4%) * | 244,689 (55.2%) | 6,245 (51.9%) * | 191,641 (54.7%) |
| **Age (years)** | 65.2 (50 – 83) * | 67.9 (48 – 86) | 65.8 (50 – 83) * | 68.3 (50 – 86) |
| **Ethnicity** |  |  |  |  |
| **White** | 14,588 (89.4%) * | 418,342 (94.3%) | 12,031 (100%) | 350,410 (100%) |
| **Other** | 1,737 (10.6%) * | 25,237 (5.7%) | 0 (0%) | 0 (0%) |
| **BMI (kg/m<sup>2</sup>)** | 28.4 (5.1) * | 27.3 (4.7) | 28.3 (5.1) * | 27.3 (4.7) |

Age is shown as mean (range). (Note that although the overall age range was 48-86, only seven total subjects, or < 0.002%, had ages outside the range 50-83.) BMI is shown as mean (SD). \* indicates different from control population with $p < 0.00001$ .

We performed logistic regressions against the number (0, 1, or 2) of adrenal-permissive *HSD3B1* 1245C alleles and calculated odds ratios for the outcome of being identified as a COVID case. We analyzed men and women of all ages together as well as men and women separately, and to determine whether there was an association specific to those of advanced age, we further broke these groups down according to age below 60, between 60 and 69, and 70 or above. The results (**Fig. 2A**) show that when looking at all ages together there was no apparent association between genotype and COVID susceptibility, but in women of age 70 or above, there was a statistically strong association between adrenal-permissive genotype and odds of identified COVID infection (odds ratio = 1.11 per C allele, 95% confidence interval = 1.04 – 1.17, p = 0.00054). In subjects of age below 60, there was a trend toward modestly reduced odds of COVID infection with adrenal-permissive genotype (OR = 0.96, 95% CI = 0.92 – 1.00, p = 0.079) driven by similar trends in each sex. Similar results were obtained in the analysis limited to the UK Biobank’s Caucasian cohort (**Fig. S2A**^35^). For women of age 70 or above, the odds ratio of 1.11 (95% CI = 1.04 – 1.18, p = 0.0013) matched that from the overall population analysis. In Caucasian subjects below age 60 the association between adrenal-permissive genotype and reduced COVID odds was stronger than in the overall population analysis (OR = 0.94, 95% CI = 0.89 – 0.98, p = 0.010). Additionally, in women of age 70 or above, there was a step-wise increase in COVID rates with increasing copies of the C allele (Caucasian cohort: AA genotype 936 cases/44,617 subjects = 2.10%, AC genotype 992/42,749 = 2.32%, CC genotype 262/10,210 = 2.57%; all races: AA genotype 1,250/55,833 = 2.24%, AC genotype 1,226/51,733 = 2.37%, CC genotype 327/12,305 = 2.66%; see **Table S1**^35^ for full breakdowns by sex and age group), suggesting that a second copy of the adrenal-permissive C allele confers additional risk above a single copy.

**Figure 2.**
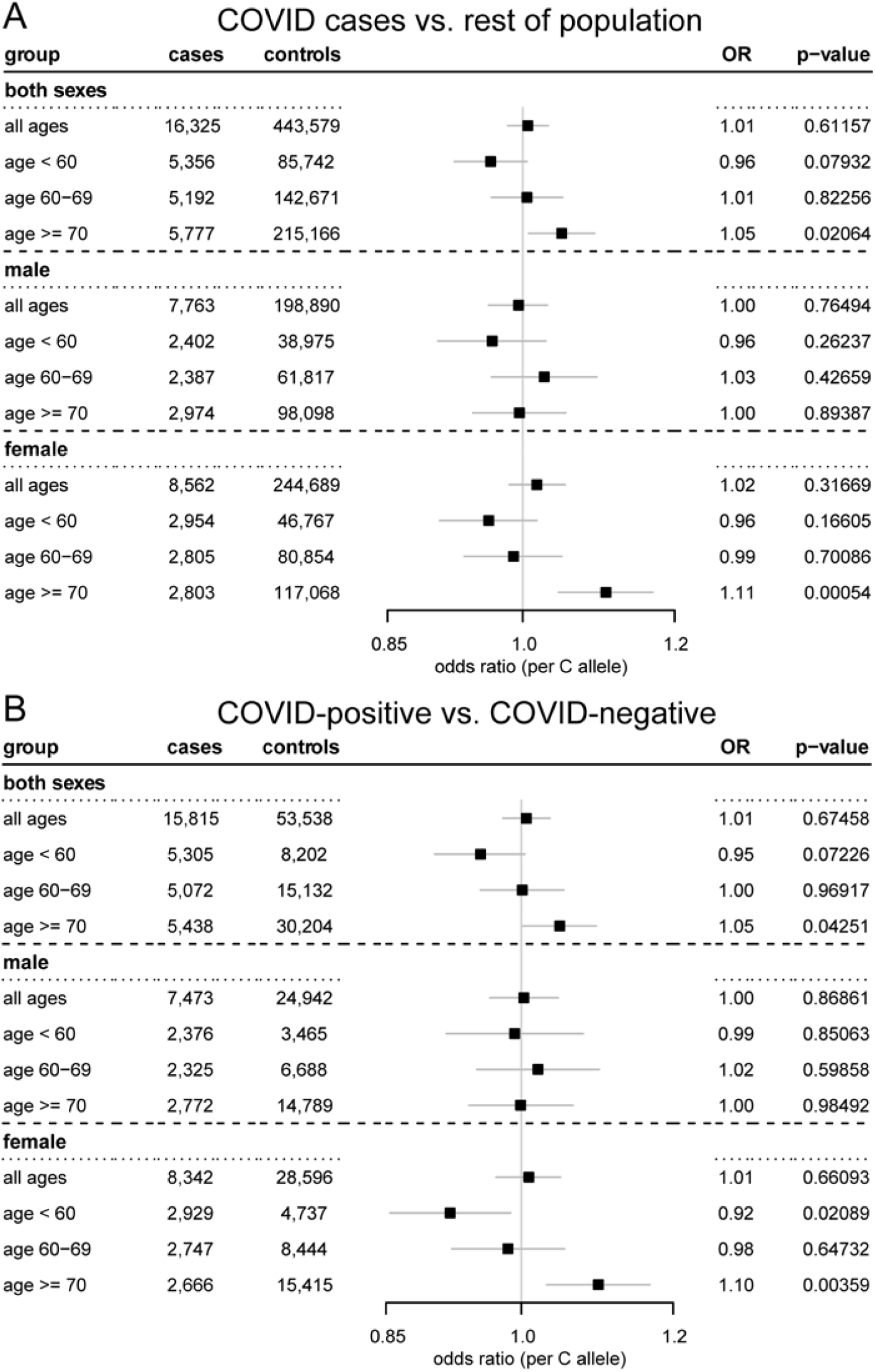
In older women, the adrenal-permissive *HSD3B1*(1245C) allele is associated with increased odds of COVID. Odds ratios (per C allele) and 95% confidence intervals for COVID in different age and sex breakdowns of the UK Biobank. **(A)** Subjects who were identified as COVID cases by positive test or ICD10 diagnosis code during inpatient hospital episode (or both) vs. all other UK Biobank subjects who were alive at the beginning of 2020. **(B)** Subjects who tested positive for COVID vs. subjects with solely negative COVID test results. Results were adjusted for sex, age, BMI, and the first ten genetic principal components (regressions including both sexes) or for age, BMI, and the first ten genetic principal components (regressions limited to one sex).

For an alternate approach to assaying COVID susceptibility, we also calculated odds ratios for testing positive for COVID among only those subjects with COVID test results, i.e., a comparison between those who tested positive and those who tested negative. This analysis yielded similar results (**Fig. 2B, Fig. S2B**^35^) to the comparison between COVID cases and the rest of the population. Notably, in women of age 70 or above there were again associations between adrenal-permissive genotype and COVID when looking within the entire subject pool (OR = 1.10, 95% CI = 1.03 – 1.17, p = 0.0036) or when limiting the analysis to the Caucasian cohort (OR = 1.09, 95% CI = 1.02 – 1.17, p = 0.011).

To test whether the association in women of elevated age might be due to selecting a cutoff age that happened to create the strongest result, we systematically varied the lower cutoff age for inclusion in the regression analysis from 50 through 80 and calculated the odds ratios and confidence intervals by year of cutoff age. **Fig. 3** shows the results for women: for each year of age, the odds ratio and confidence interval for COVID cases among women of that age or older are shown. Note that a vertical slice through the graph at age 70 is equivalent to the “female, age ≥ 70” row in Fig. 2A. For any lower cutoff age from 59 through 80, the result that an association between the C allele and women of at least that age having elevated COVID odds, with a 95% confidence interval not overlapping an odds ratio of 1.0, would hold true. Because results similar to our initial cutoff were obtained over a wide range of cutoff ages, the association in women of elevated age is not the result of having selected a specific age. As the cutoff age increases, the odds ratio tends to become larger; at the same time, the confidence interval grows wider as the sample size decreases (e.g., at a cutoff age of 70, subjects of age 70 and up are included; at a cutoff age of 80, only subjects of age 80 and up are included). For the COVID positivity among COVID tested women odds ratios, a cutoff age ranging anywhere from 64 through 76 would result in the 95% confidence interval not overlapping 1.0, with the lower bound of the confidence interval only crossing 1.0 at some cutoff ages above 76 due to decreasing sample sizes (**Fig. S3**^35^), further demonstrating that the association with genotype in women of elevated age is not an artifact of having selected a specific cutoff age.

**Figure 3.**
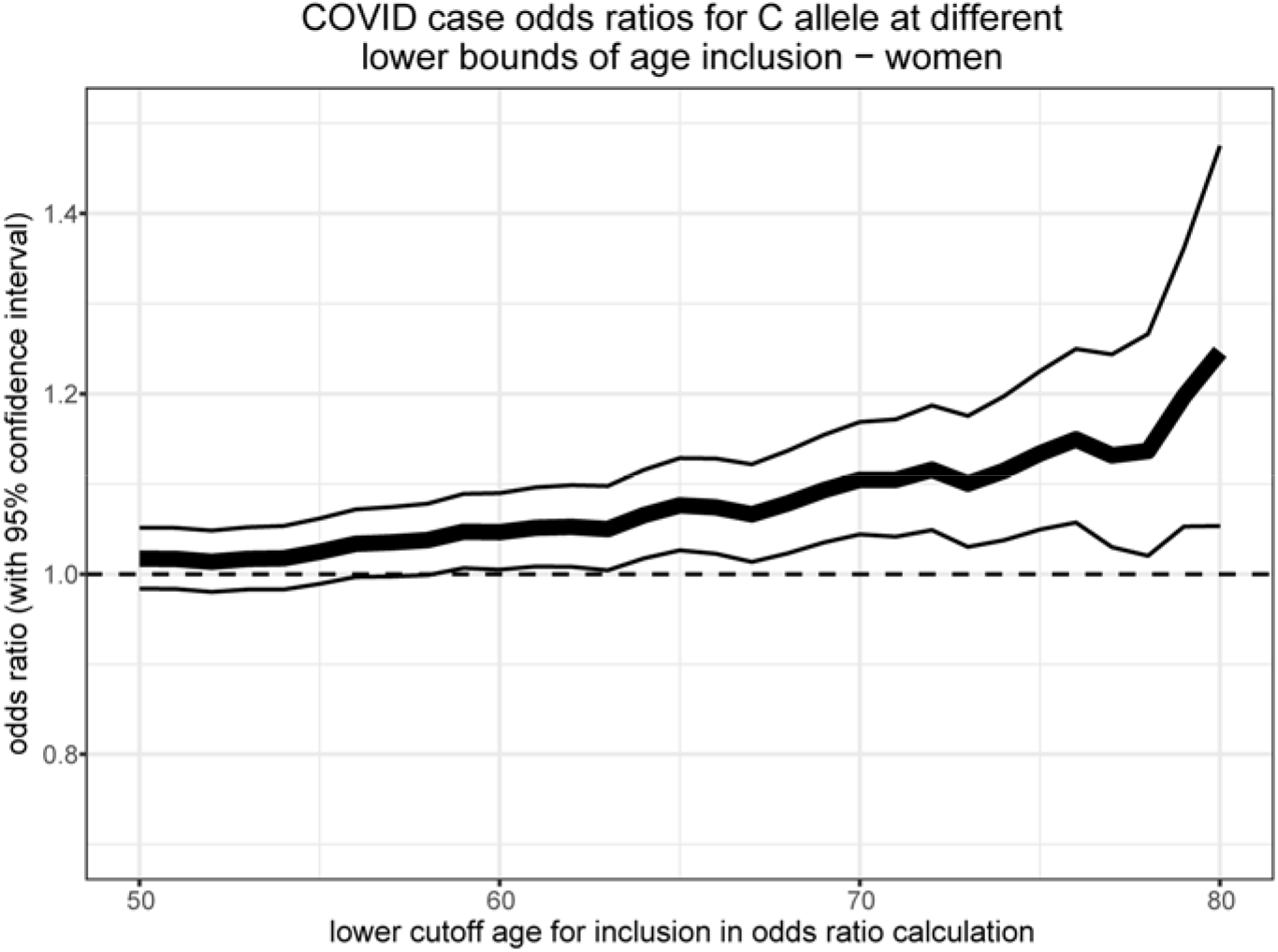
Adrenal-permissive genotype associates with higher COVID odds in older women regardless of the lower cutoff age for inclusion in the analysis. Odds ratios per C allele (thick line) and 95% confidence intervals (thin lines) for being identified as a COVID case in women at least [cutoff age] years old, by cutoff age. For each year of age, the odds ratio and confidence interval in women that age or older were calculated. Note that the result at a lower cutoff age of 70 is the same as the “female, age ≥ 70” result in **Fig. 2A**.

The UK’s COVID testing strategy was not constant over the course of the investigation period, due to especially limited resources at the beginning of the pandemic. According to the Oxford Covid-19 Government Response Tracker, prior to May 18, 2020, access to testing was restricted to people meeting specific criteria, whereas from that point on it was available to anyone showing symptoms^36^. To assess whether the changing availability of testing affected the apparent association between adrenal-permissive genotype and COVID susceptibility, we repeated the analyses but split into two time periods: first restricting results to testing and diagnoses that occurred prior to May 18, 2020, and then restricting results to May 18, 2020, onward. Although the sample sizes were much smaller for the first period, associations between both COVID case identification and test positivity were observed in women ages 70 and above in both periods (odds ratios for COVID case identification: early pandemic 1.18 (95% CI 1.00 – 1.38, p = 0.0456), subsequent period 1.09 (95% CI 1.03 – 1.16, p = 0.00415); odds ratios for COVID test positivity: early pandemic 1.27 (95% CI 1.03 – 1.58, p = 0.0285), subsequent period 1.08 (95% CI 1.02 – 1.16, p = 0.0153). There was some variability of odds ratios, but for all age and sex breakdowns, there was substantial overlap in the confidence intervals across the two time periods, suggesting that changing availability of testing had little effect on results (**Fig. S4**^35^).

To test for an association between *HSD3B1* genotype and COVID outcomes, we analyzed the UK Biobank’s death records and calculated the odds ratios for death by number of C alleles among subjects who had COVID. Here we analyzed men and women of all ages together, men and women separately, and age groups below 70 and 70 or above, rather than separating ages below 60 from 60 – 69, due to small numbers of deaths below age 60. This analysis did not show an association between genotype and COVID outcomes in the whole population (**Fig. 4**) or Caucasian cohort (**Fig. S5**^35^), although the numbers of events for this analysis were considerably smaller than for the COVID cases and positivity analyses, especially for the lower age ranges. The mortality analysis included deaths with any listed cause of death subsequent to identification as a COVID case; restricting the included deaths to those with COVID listed as a cause of death similarly did not reveal associations with genotype (not shown).

**Figure 4.**
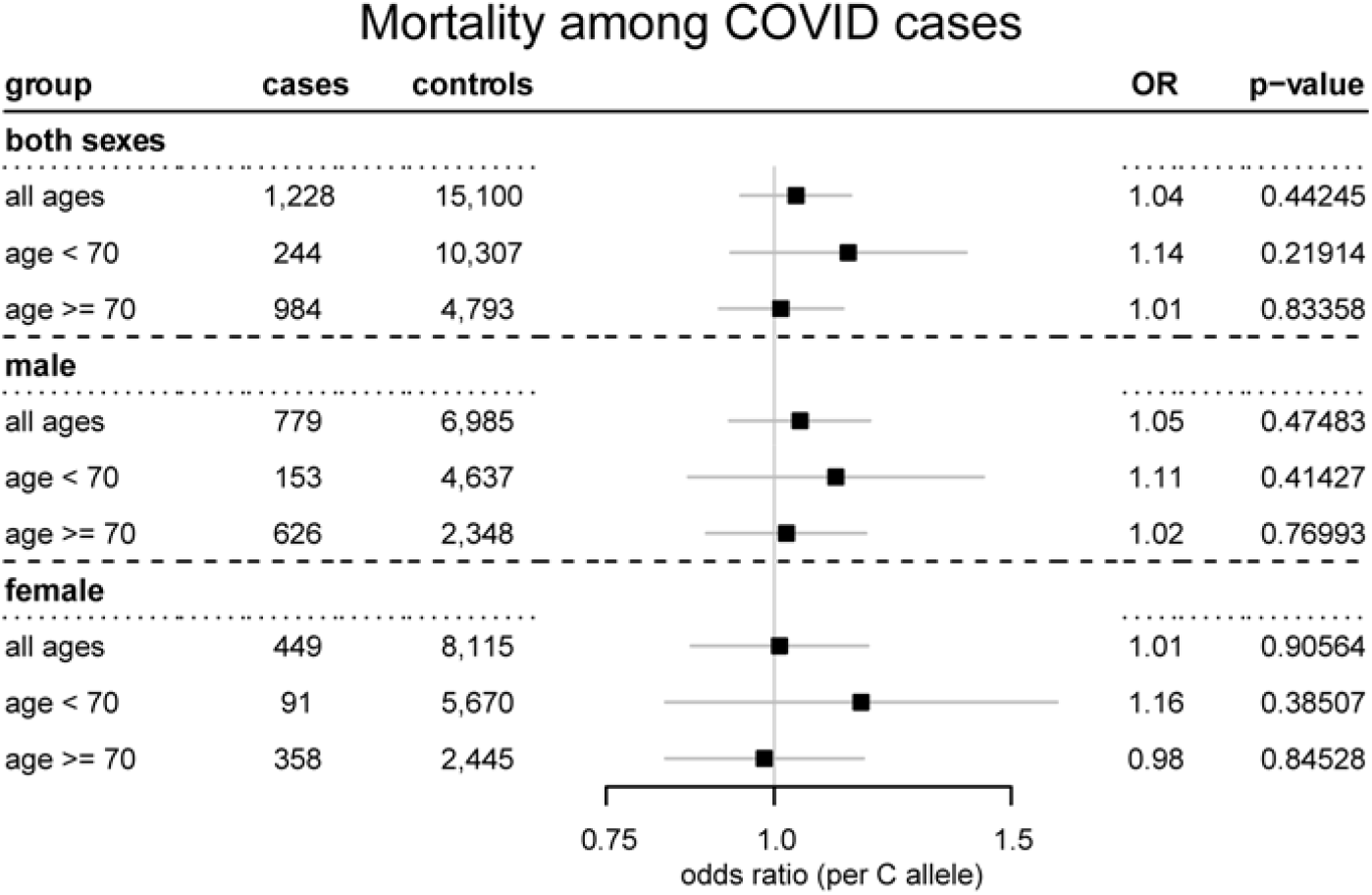
*HSD3B1* genotype does not appear to be associated with COVID outcomes. Odds ratios (per C allele) and 95% confidence intervals for death among subjects with identified COVID cases in different age and sex breakdowns of the UK Biobank. Results were adjusted for sex, age, BMI, and the first ten genetic principal components (regressions including both sexes) or for age, BMI, self-reported ethnicity, and the first ten genetic principal components (regressions limited to one sex).

To attempt to explore the mechanistic basis for the associations we found, we interrogated whether there were associations between *HSD3B1* genotype and the levels in circulation of the steroids for which data are available in the UK Biobank: testosterone and estradiol. Concentrations of these steroids were measured in serum samples collected when participants enrolled in the study in 2006-2010. For testosterone, we performed analyses in men and women separately, as well as specifically in women who were premenopausal or postmenopausal upon entering the study. For estradiol, >90% of men and of postmenopausal women had levels below the bottom of the reportable range (175 pM), so we only analyzed the data from premenopausal women. We did not find that testosterone or estradiol levels differed by genotype (**Fig. S6**^35^), which is consistent with the results from our previous study in which *HSD3B1* genotype was found to be associated with response to glucocorticoid treatment in asthma but circulating testosterone levels were not found to differ by genotype^21^.

Lastly, because we observed that the COVID odds ratios for women of elevated age tended to increase with an increasing lower cutoff age, we examined the relationship between subject age and frequencies of adrenal-permissive and adrenal-restrictive genotypes among subjects identified as having had COVID. We graphed the adrenal-permissive (1245C) allele frequencies of Caucasian cohort subjects who had COVID by year of age and found that in women, there was a linear relationship between age and C allele frequency (adjusted R^2^ = 0.37, p = 7.87e-05 [regression weighted by number of cases per year of age], **Fig. 5A**), whereas in men, there was no such relationship (adjusted R^2^ = -0.01, p = 0.437, **Fig. 5B**). The data for these graphs were limited to the Caucasian cohort because they show raw allele frequencies rather than results of ethnicity-adjusted regression analyses, and overrepresentation of Caucasians at older ages would confound the results and make the relationships appear even stronger if the entire population were included. This relationship in women was specific to COVID cases and was not present for the overall population (adjusted R^2^ = 0.01, p = 0.241 for all Caucasian women alive in 2020). To more clearly visualize how the frequencies of adrenal-permissive vs. adrenal-restrictive genotypes varied with age, we also graphed the adrenal-permissive (AC or CC) and adrenal-restrictive (AA) frequencies of Caucasian cohort subjects who had COVID by a moving window of ages, illustrating that in women, adrenal-permissive genotype frequencies steadily increased with increasing age (**Fig. 5C**), whereas in men, there was no clear trend with age (**Fig. 5D**). In women, at the lower end of the age range the adrenal-permissive genotype frequency of COVID cases is marginally lower than the overall population frequency; at the upper end of the age range, it is substantially higher. These results imply that the association between adrenal-permissive genotype and COVID susceptibility becomes increasingly strong with increasing age in women.

**Figure 5.**
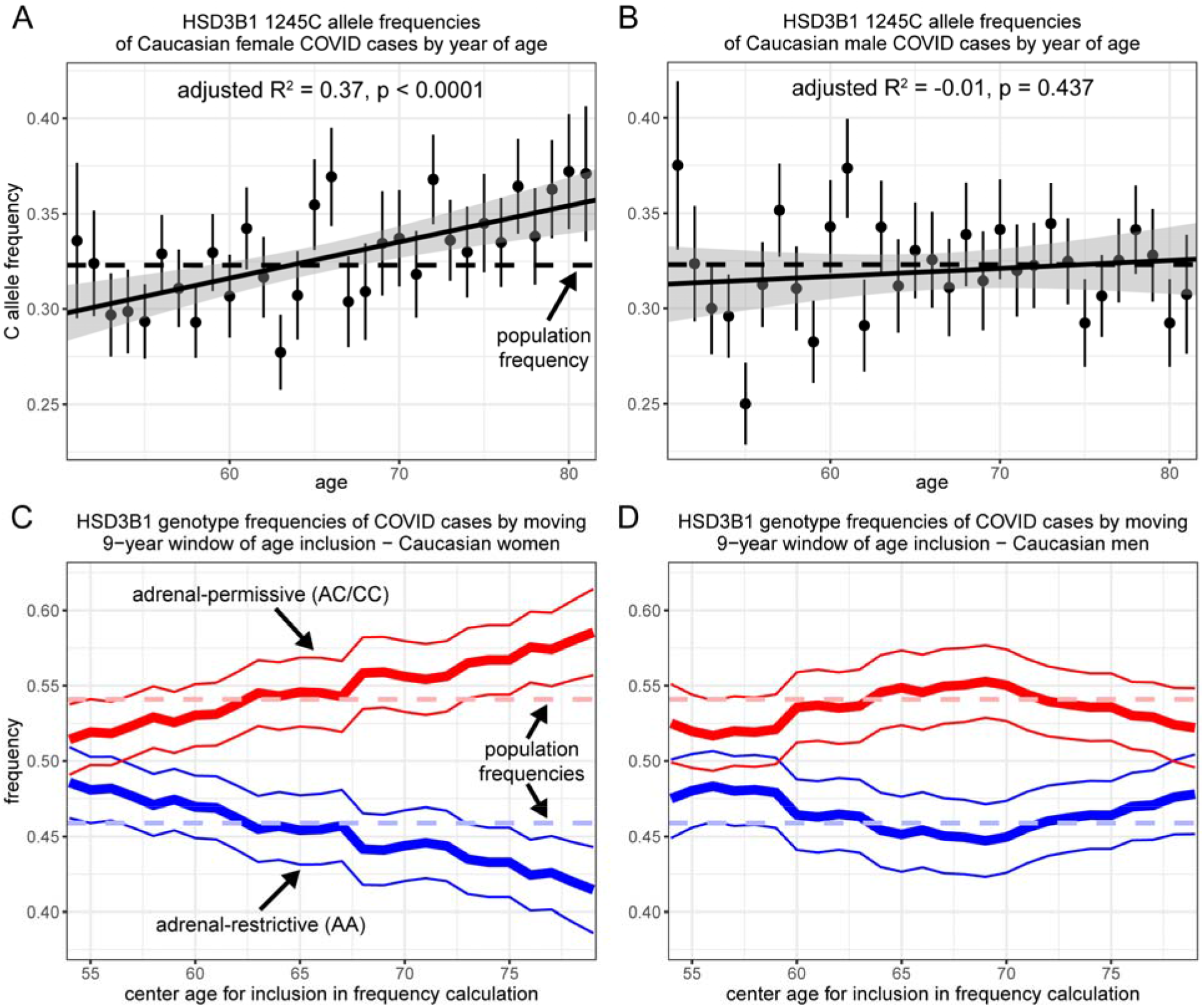
There is a positive linear relationship between age and adrenal-permissive genotype frequency in women identified as COVID cases. **(A-B)** The C allele frequencies (± SEM) for female **(A)** and male **(B)** subjects in the UK Biobank Caucasian cohort who had COVID for each year of age, along with the lines of best fit and 95% confidence intervals from weighted linear regressions of allele frequency ∼ year of age weighted by number of cases for each year of age. The dashed horizontal lines indicate the C allele frequencies of the overall cohorts. Ages 50, 82, and 83 had just 11, 17, and 4 cases, respectively, out of a total of 6,245 cases for women, and 11, 36, and 0 cases out of a total of 5,786 cases for men, and are not shown in the graphs but were included in the weighted linear regressions. **(C-D)** The smoothed frequencies of adrenal-permissive (AC or CC) and adrenal-restrictive (AA) genotypes among female **(C)** and male **(D)** subjects in the UK Biobank Caucasian cohort who had COVID by moving 9-year windows of age inclusion. Thick lines indicate the genotype frequencies and thin lines indicate the 95% confidence intervals. Dashed horizontal lines indicate the overall population frequencies for AC/CC (upper) and AA (lower) genotypes. For each year of age, the genotype frequencies and confidence intervals were calculated for the group of subjects identified as COVID cases and with age equal to the given year ± 4. For example, the data points for age 70 include subjects from ages 66 through 74.

## Discussion

We have demonstrated that, in the UK Biobank, there is an association between possessing the adrenal-permissive *HSD3B1* 1245C allele and being identified as having COVID-19 in women of elevated age, i.e., around 70 and older. This association exists whether comparing those subjects who were identified as COVID cases to the rest of the population (**Fig. 2A, Fig. S2A**) or comparing those subjects who tested positive for COVID to those who tested negative (**Fig. 2B, Fig. S2B**). This association in older women is not an artifact of having selected a specific cutoff age, but rather is robust to using a wide range of cutoff ages (**Fig. 3, Fig. S3**). On the other hand, we did not detect an association between *HSD3B1* genotype and COVID outcomes (**Fig. 4, Fig. S4**). Lastly, we found that the pool of women with COVID became increasingly enriched for adrenal-permissive genotypes with increasing age (**Fig. 5**), suggesting that adrenal-permissive steroid biosynthesis may have an increasing effect on immune response as women age. Although statistically well supported, the associations we observed did not have enormous effect sizes (odds ratio ∼1.1 per risk allele in women of advanced age), so it does not appear that *HSD3B1* genotype is a strong determinant of an individual’s COVID risk; rather, the results are of interest for the potential implications regarding the role of steroids in the immune system.

Previously reported phenotypic associations with adrenal-restrictive vs. adrenal-permissive *HSD3B1* genotype, in both prostate cancer and asthma, occur in contexts of reduced circulating androgen levels. In men, the pool of gonadally produced androgens is vastly larger than in women, so the relative contribution of adrenally produced androgens is greater in women. Furthermore, in postmenopausal women, the adrenals are the sole source of sex steroids, whereas men continue to produce androgens gonadally throughout life^37,38^. As menopause typically occurs between ages 45 and 55 and over 90% of UK Biobank subjects are greater than 55 years old, the female cohort is very predominantly postmenopausal. Additionally, circulating levels of the adrenally produced precursor DHEA that is a 3βHSD1 substrate in the pathway to potent androgen production decrease with increasing age^39,40^. Considering these points together, one could speculate that the adrenal-permissive genotype would have a more pronounced physiologic effect in postmenopausal women that would become increasingly strong with increasing age, which is in line with our results showing that as women’s ages increase, the association between possessing the adrenal-permissive C allele and being identified as having COVID becomes increasingly strong (**Fig. 5**). As COVID outcomes are by far the worst in elderly patients, the existence of a germline steroid-metabolizing enzyme variant that is associated with heightened susceptibility to COVID in older women would be of physiologic and potentially pharmacologic importance.

Although it is well established that men are at greater risk of severe disease and death from COVID than women, existing data make it less clear whether there is any disparity between men and women in susceptibility to infection. A large meta-analysis covering studies from around the globe reported that the overall proportion of male COVID cases was exactly half^41^, and The Sex, Gender, and COVID-19 Project, which tracks all sex-disaggregated COVID cases reported worldwide, similarly shows as of April 2021 that male cases account for 49.2% of the 73,793,217 sex-disaggregated cases globally^42^. These results indicate that men and women are infected at the same rate, but other studies suggest that the lack of observed sex discordance is an artifact of all ages being grouped together. Multiple studies have reported that case rates are higher in women from roughly ages 20 to 60 with a reversal to being higher in men above age 60^43-45^. This has been attributed to women being overrepresented among high-exposure occupations such as health care workers^43,45^. Evidence for sex hormone mediated modulation of immune responses, the precise mechanisms of which are still the subject of much ongoing research^10,13^, raises the question of whether pre vs. postmenopausal hormone environments could also be a factor. Our results, from a cohort predominantly above age 60, are in agreement with this observed trend, as COVID case rates in the UK Biobank subject population are higher among men than women (**Table 1**), with this difference being driven by the 60 and up portion of the population (**Table S1**). Taken together, this body of evidence indicates that men, or at the very least older men, may in fact be more susceptible to COVID infection than women, although disentangling the contributions of gender-based differences in occupational risk of exposure, gender-based differences in compliance with protective measures such as social distancing and mask-wearing^46^, and sex-based differences in biological susceptibility to infection is difficult. An additional caveat is the question of whether differences are being measured in total infection rates or in rates of symptomatic infection. Nonetheless, our finding that women of elevated age who inherited the variant form of 3βHSD1 conferring increased androgen synthesis have higher COVID rates is consistent with androgen-mediated immune suppression being a contributing factor to the higher COVID rates in older men vs. women.

Whereas our results suggest that increased androgen-mediated immune suppression in older women of adrenal-permissive genotype could lead to increased susceptibility to COVID infection, we did not find evidence for a similar effect in outcomes among COVID patients (**Fig. 4, Fig. S4**). It was shown in a large, randomized trial that treatment with the glucocorticoid dexamethasone strongly reduced mortality in COVID patients on ventilation but not in patients receiving no respiratory support^47^, and dexamethasone has since become a mainstay treatment for hospitalized COVID patients with severe disease. Taken together with the findings we report here and our previous finding that *HSD3B1* genotype affects response to glucocorticoid treatment in asthma with the adrenal-permissive 1245C allele associated with reduced airway inflammation^21^, this raises the question of whether *HSD3B1* genotype could affect response to dexamethasone in COVID patients, and whether adrenal-permissive genotype could be beneficial in such patients, as it is in the asthma setting. This question could not be addressed using the data available in the UK Biobank and should be the subject of future studies. The differential effects of dexamethasone in COVID patients who were or were not receiving respiratory support also highlight how the results of steroid-induced immune suppression could be context-dependent: immune suppression could make it harder to ward off an initial infection, but could be beneficial in a patient with an advanced infection and out-of-control inflammatory response.

The finding of increased COVID rates in older women with adrenal-permissive genotype has the strongest statistical support of our results, but we also found some evidence that in younger (below age 60) subjects of both sexes, adrenal-permissive genotype was associated with modestly reduced COVID rates, i.e., an association in the opposite direction (**Fig. 2, Fig. S2**). The statistical support for this finding is not nearly as strong, so this result should be interpreted with caution and discussion of its implications should be regarded with this caution in mind. The products of 3βHSD1-catalyzed reactions include immediate precursors of both androgens and estrogens, and estrogens have been reported to bolster immune response^9-11^, including in a mouse study of the SARS-CoV-1 virus that caused the 2003 SARS outbreak^48^ and is closely related to the SARS-CoV-2 virus that causes COVID. If the modestly reduced COVID rates in younger subjects with adrenal-permissive genotype represent a real trend, this suggests the possibility that the interplay of androgens and estrogens in regulating immune response could vary in both age and sex-dependent ways; i.e., cells expressing *HSD3B1*(1245C) could potentially have heightened production of both androgens and estrogens, the effects of which might vary with age and/or sex.

In women identified as COVID cases, we found a strong linear relationship between age and adrenal-permissive *HSD3B1*(1245C) allele frequencies (**Fig. 5**). This linear relationship covering the range of roughly 50 through 80 years of age raises the question of whether the same relationship would continue over a wider age range. It is tempting to speculate that adrenal-permissive genotype would continue to become more strongly associated with susceptibility to infection if the age range were expanded upward but that there would be a plateau rather than a continued decrease if the age range were expanded downward to premenopausal ages. In postmenopausal women, the adrenals are the sole source of sex steroids, and DHEA levels continue to decline with increasing age. By contrast, in premenopausal women, the adrenally produced sex steroid pool is in addition to the gonadally produced pool. However, it is not possible to address this question using the UK Biobank; women’s menopausal statuses when they enrolled in the study are available, but not menopausal statuses as of 2020. In the under 60 age range contained in the UK Biobank (i.e., 50-59), most but not all women would be postmenopausal, and menopausal statuses of individual women not previously identified as having undergone menopause cannot be ascertained. To address the question of whether the association is different in pre vs. postmenopausal women, a subject population with sufficient sample sizes over a wider range of ages is required.

Our study did not identify the mechanistic basis for the observed association between adrenal-permissive genotype and COVID susceptibility. We analyzed circulating testosterone levels in men and women as well as circulating estradiol levels in premenopausal women and did not find that levels differed by genotype. This is consistent with a previous study that did not find differences in testosterone levels by genotype in male or female asthma patients^21^; the results we report here come in a general population with much larger sample sizes. Despite the apparent lack of an appreciable effect of *HSD3B1* genotype on circulating steroid levels, clear phenotypes with *HSD3B1* genotype have been observed in two asthma cohorts^21^ and at least eight prostate cancer cohorts^49^, suggesting that the effects of *HSD3B1* genotype are mediated largely independent of circulating sex steroid levels. Steroid levels in tissues can differ profoundly from those in circulation^50-52^, so *HSD3B1* genotype could potentially affect concentrations at the cell or tissue level without measurably affecting concentrations in circulation, which should be further studied. Other measures such as circulating cortisol levels and inflammatory cytokines are also important to analyze, but these data were not available in the UK Biobank.

The results we report here should be validated in an independent cohort. In any validation attempt, it is critical to disaggregate the results by both sex and age range rather than simply grouping all subjects together, as well as to obtain sufficient sample sizes in the demographic (older women) in which we observed the strongest association. If this validation is forthcoming, it would be an important finding in both the field of COVID research as well as in the broader area of the role of sex hormones in immune responses. Importantly, the existence of a differential immune response based on differential androgen metabolism occurring specifically within one sex (i.e., women) could help shed light on the role of androgens in immune responses in general. Comparisons between men (higher androgen levels) and women (lower androgen levels) are confounded by numerous other physiologic differences between the sexes; in a comparison between women with adrenal-permissive and women with adrenal-restrictive androgen synthesis, these confounding factors are absent. The mechanisms underlying associations between immune responses and inherited adrenal androgen metabolism by way of *HSD3B1* genotype are subjects for future study.

## Data Availability

Data available per regulations of UK Biobank.

## Author contributions

Jeffrey M. McManus: conceptualization, methodology, investigation, writing – original draft, visualization; Navin Sabharwal: methodology, investigation, data curation; Peter Bazeley: methodology, data curation; Nima Sharifi: conceptualization, writing – review & editing, supervision, funding acquisition

## Data and materials availability

Data are regulated by UK Biobank’s policies on access.

