## Supplemental materials for "Inheritance of a common androgen synthesis variant allele is associated with female COVID susceptibility in UK Biobank"

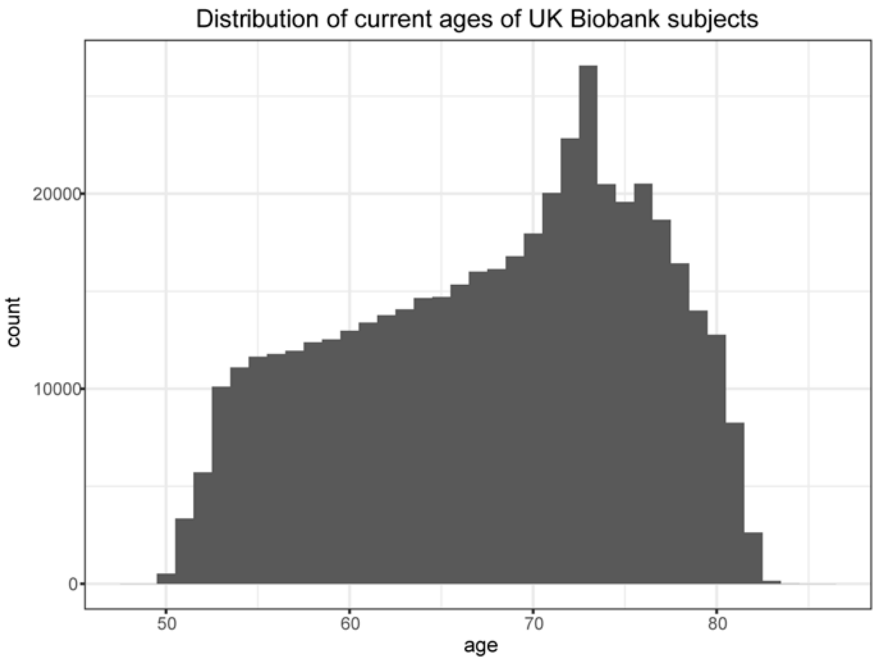


**Figure S1. Distribution of current ages of UK Biobank subjects included in analyses.** Subjects included in analyses were subjects with *HSD3B1* genotype available and who were alive as of the beginning of 2020. Current ages were determined using subjects' birth years and months and were calculated as of September 2020, the midpoint of the COVID results.

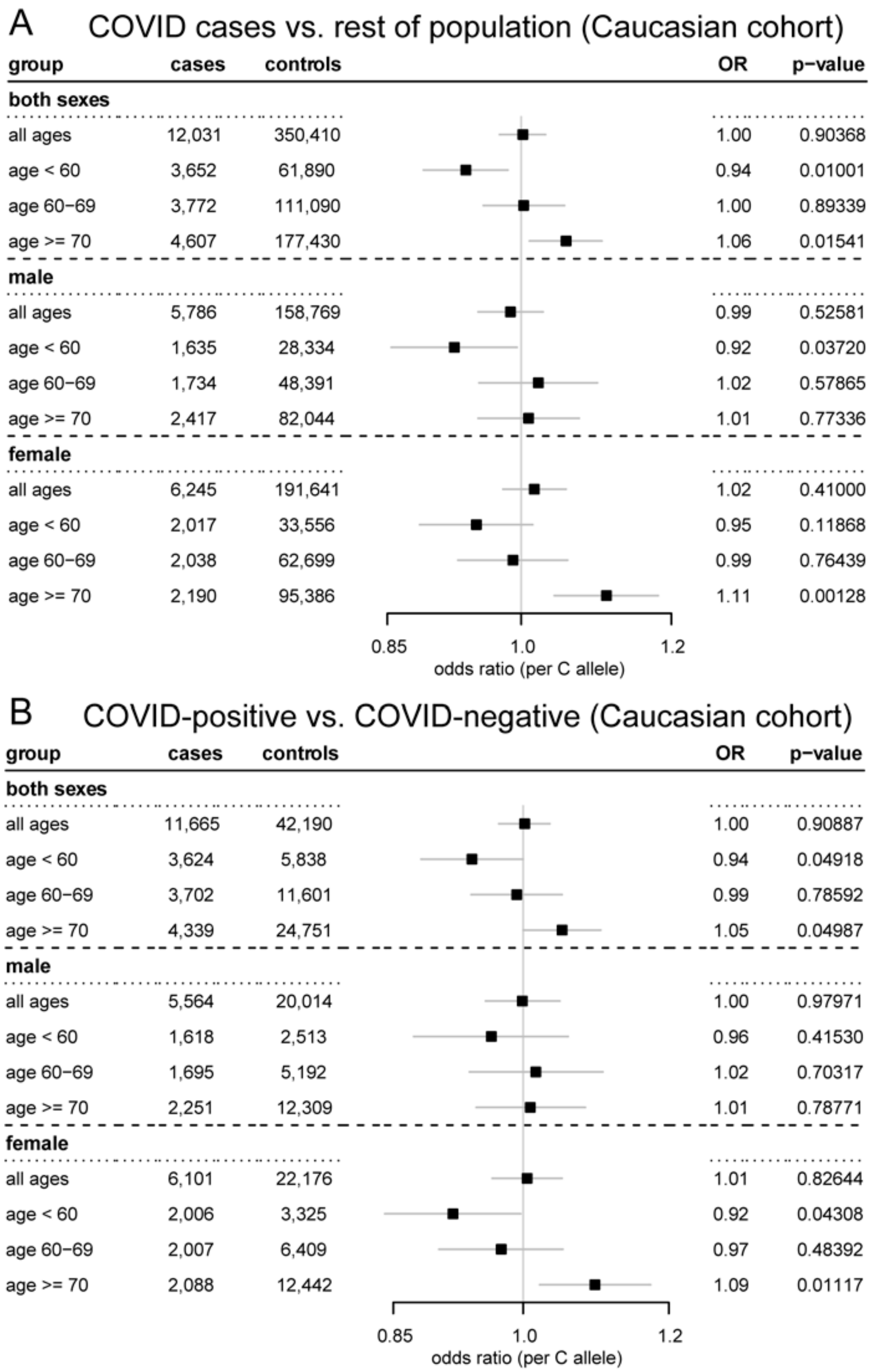


**Figure S2. In older women of the Caucasian cohort, the adrenal-permissive *HSD3B1*(1245C) allele is associated with increased odds of COVID.** Odds ratios (per C allele) and 95% confidence intervals for COVID in different age and sex breakdowns of the UK Biobank's Caucasian cohort as defined by the UK Biobank. **(A)** Subjects who were identified as COVID cases by positive test or ICD10 diagnosis code during inpatient hospital episode (or both) vs. all other UK Biobank Caucasian subjects who were alive at the beginning of 2020. **(B)** Subjects who tested positive for COVID vs. subjects with solely negative COVID test results. Results were adjusted for sex, age, BMI, and the first ten genetic principal components (regressions including both sexes) or for age, BMI, and the first ten genetic principal components (regressions limited to one sex).

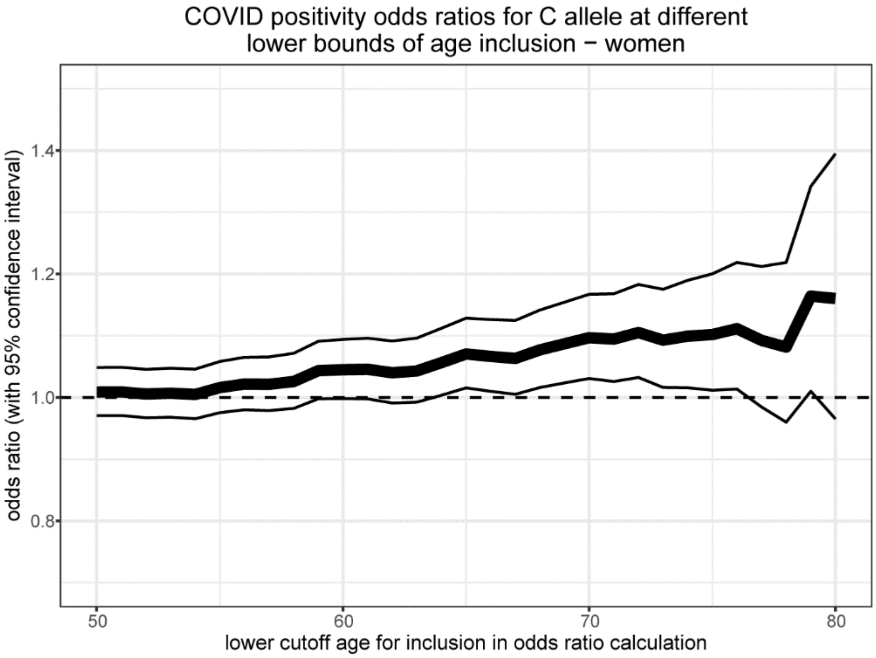


**Figure S3. Adrenal-permissive genotype associates with higher COVID positivity in older women regardless of the lower cutoff age for inclusion in the analysis.** Odds ratios per C allele (thick line) and 95% confidence intervals (thin lines) for testing positive for COVID among women at least [cutoff age] years old who were tested for COVID, by cutoff age. For each year of age, the odds ratio and confidence interval in women that age or older were calculated. Note that the result at a lower cutoff age of 70 is the same as the “female, age ≥ 70” result in **Fig. 2B**.

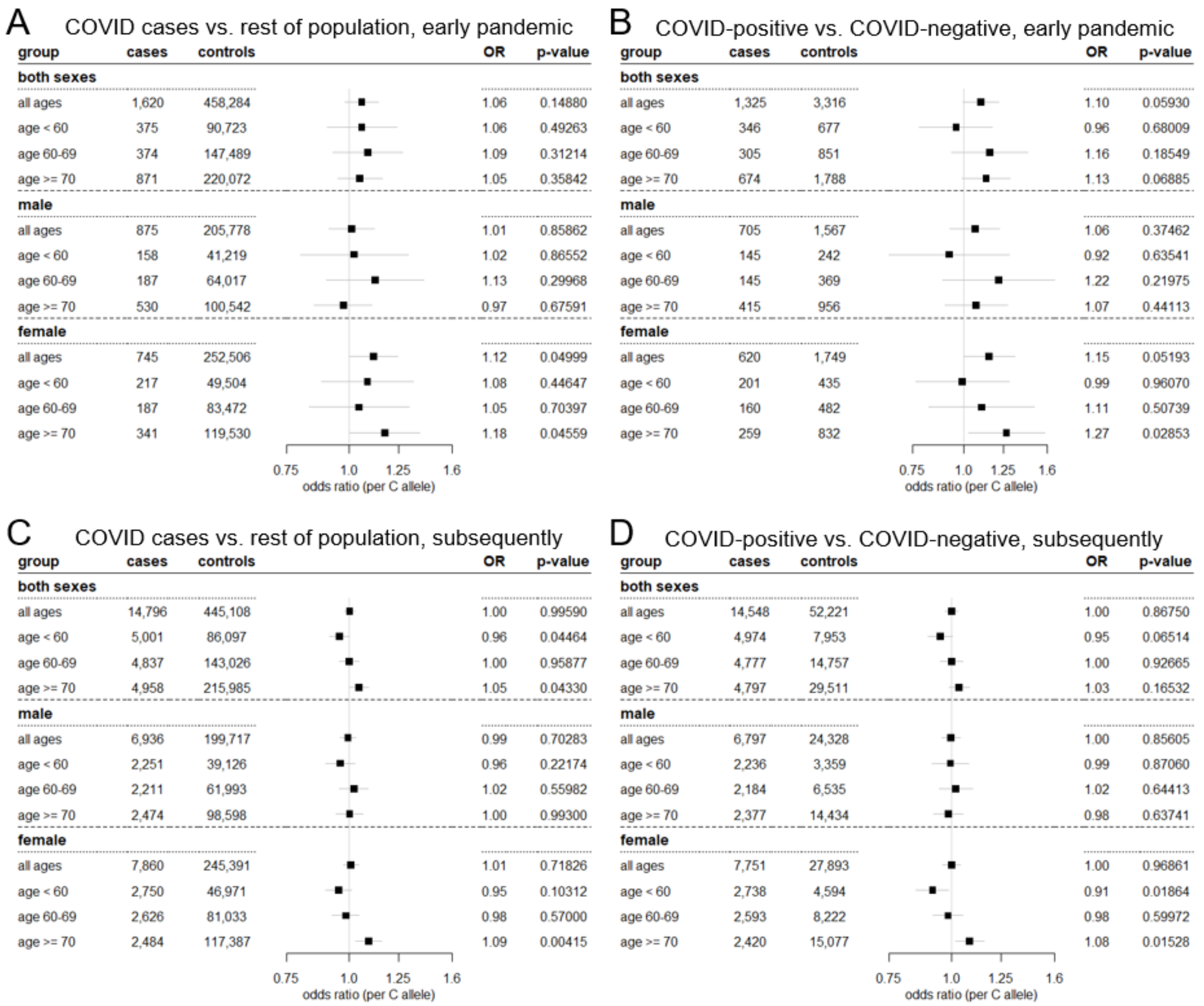


**Figure S4. Associations by period of pandemic according to UK COVID testing strategy.** Same results as in **Fig. 2** but with COVID testing and diagnoses restricted to one of two time periods: pre-May 18, 2020, when testing availability was limited to people meeting specific criteria, and subsequently, when testing was available to anyone with symptoms. **(A)** Odds ratios for COVID cases vs. rest of population, pre-May 18, 2020. **(B)** Odds ratios for positive vs. solely negative COVID testing, pre-May 18, 2020. **(C)** Odds ratios for COVID cases vs. rest of population, May 18, 2020, and onward. **(D)** Odds ratios for positive vs. solely negative COVID testing, May 18, 2020, and onward.

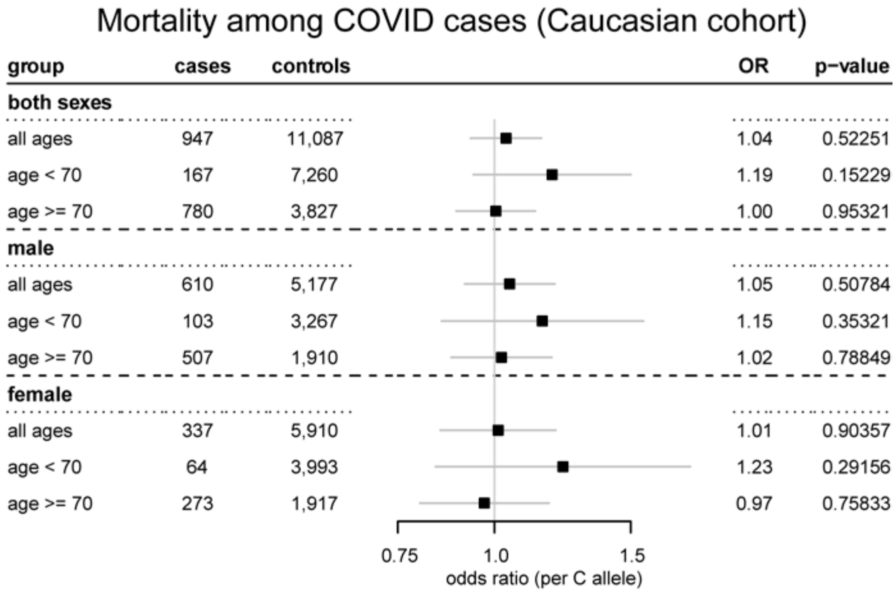


**Figure S5. *HSD3B1* genotype does not appear to be associated with COVID outcomes in analysis restricted to Caucasian cohort.** Odds ratios (per C allele) and 95% confidence intervals for death among subjects with identified COVID cases in different age and sex breakdowns of the UK Biobank Caucasian cohort. Results were adjusted for sex, age, BMI, and the first ten genetic principal components (regressions including both sexes) or for age, BMI, and the first ten genetic principal components (regressions limited to one sex).

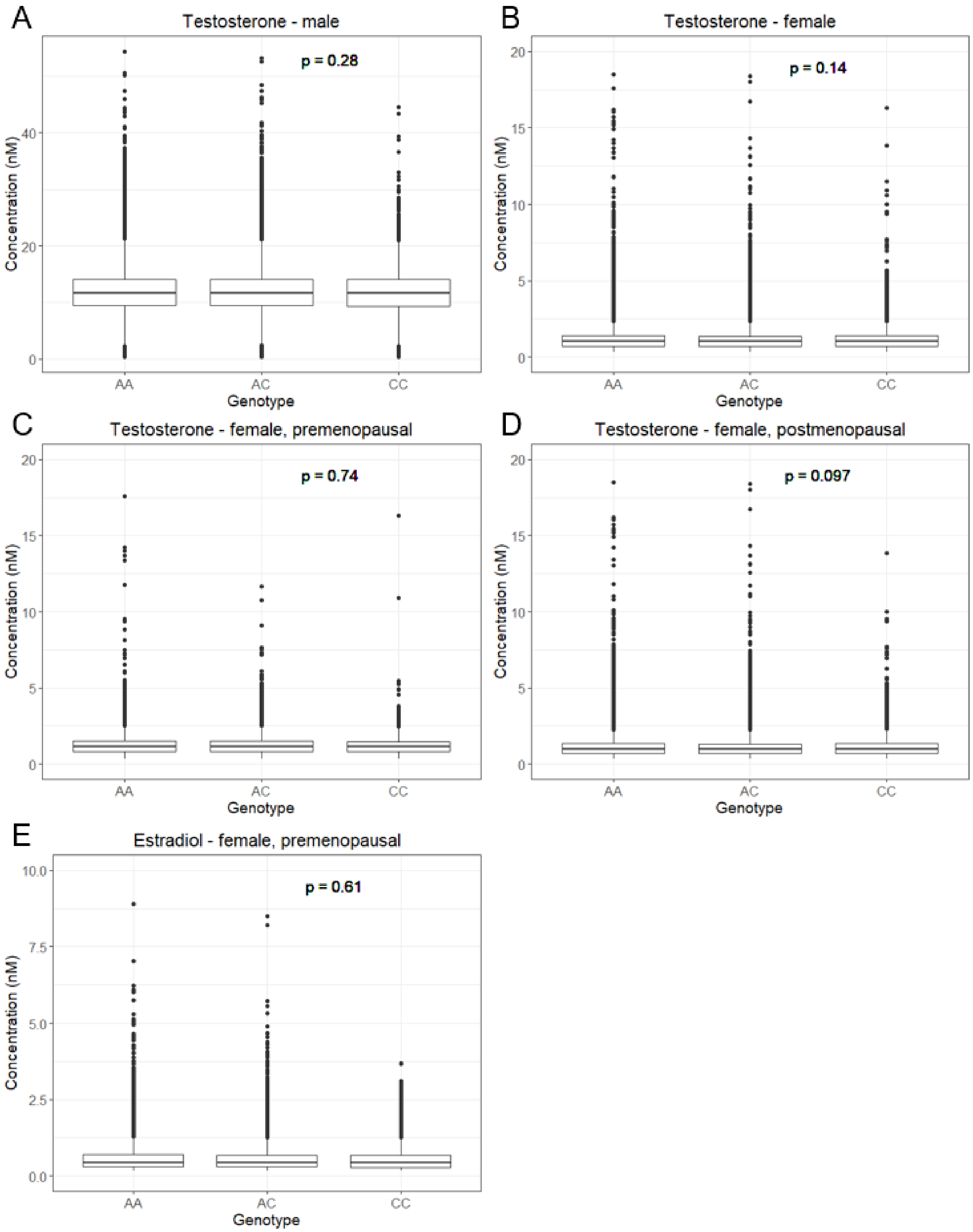


**Figure S6. Circulating testosterone and estradiol levels are similar across *HSD3B1* genotypes.** Box-and-whisker plots of testosterone and estradiol concentrations by genotype in the UK Biobank Caucasian cohort measured in blood samples obtained when subjects enrolled. Center lines indicate median values, boxes indicate first quartile to third quartile range, whiskers indicate values up to 1.5 times interquartile range, and points indicate outlying values. **(A)** Testosterone concentrations in men (n = 77,303 AA genotype, 73,529 AC, 17,488 CC). **(B)** Testosterone concentrations in women (n = 75,388 AA, 71,833 AC, 16,988 CC; 7 outlier values above 20 nM were excluded from plot for visual clarity). **(C)** Testosterone concentrations in premenopausal women (n = 18,295 AA, 17,267 AC, 4,071 CC). **(D)** Testosterone concentrations in postmenopausal women (n = 53,765 AA, 51,479 AC, 12,136 CC; 7 outlier values above 20 nM were excluded from plot for visual clarity). **(E)** Estradiol concentrations in premenopausal women (n = 13,455 AA, 12,656 AC, 3,005 CC; 7 outlier values above 10 nM were excluded from plot for visual clarity.) Greater than 90% of men and postmenopausal women had estradiol concentrations below the reportable range, so only premenopausal women were analyzed for estradiol. P-values from Kruskal-Wallis tests are shown.

**Table S1. Breakdown of COVID case numbers and rates by *HSD3B1* genotype for each sex and age range in the UK Biobank whole population and Caucasian cohorts.**

| | | | Male | | | Female | | |
|---|---|---|---|---|---|---|---|---|
| Cohort | Age range | Genotype | COVID cases | Total subjects | COVID rate (95% CI) | COVID cases | Total subjects | COVID rate (95% CI) |
| UKBB | < 60 | AA | 1,208 | 20,075 | 6.02% (5.69 – 6.35%) | 1,501 | 24,149 | 6.22% (5.91 – 6.52%) |
| | | AC | 985 | 17,297 | 5.69% (5.35 – 6.04%) | 1,178 | 20,719 | 5.69% (5.37 – 6.00%) |
| | | CC | 209 | 4,005 | 5.22% (4.53 – 5.91%) | 275 | 4,853 | 5.67% (5.02 – 6.32%) |
| | 60-69 | AA | 1,150 | 30,314 | 3.79% (3.58 – 4.01%) | 1,366 | 39,890 | 3.42% (3.25 – 3.60%) |
| | | AC | 996 | 27,429 | 3.63% (3.41 - 3.85%) | 1,187 | 35,425 | 3.35% (3.16 – 3.54%) |
| | | CC | 241 | 6,461 | 3.73% (3.27 – 4.19%) | 252 | 8,344 | 3.02% (2.65 – 3.39%) |
| | 70+ | AA | 1,428 | 47,250 | 3.02% (2.87 – 3.18%) | 1,250 | 55,833 | 2.24% (2.12 – 2.36%) |
| | | AC | 1,232 | 43,621 | 2.82% (2.67 – 2.98%) | 1,226 | 51,733 | 2.37% (2.24 – 2.50%) |
| | | CC | 314 | 10,201 | 3.08% (2.74 – 3.41%) | 327 | 12,305 | 2.66% (2.37 – 2.94%) |
| UKBB Cauc. | < 60 | AA | 783 | 13,769 | 5.69% (5.30 – 6.07%) | 970 | 16,399 | 5.91% (5.55 – 6.28%) |
| | | AC | 705 | 13,084 | 5.39% (5.00 – 5.78%) | 845 | 15,507 | 5.45% (5.09 – 5.81%) |
| | | CC | 147 | 3,116 | 4.72% (3.97 – 5.46%) | 202 | 3,667 | 5.51% (4.77 – 6.25%) |
| | 60-69 | AA | 784 | 22,925 | 3.42% (3.18 – 3.66%) | 932 | 29,723 | 3.14% (2.94 – 3.33%) |
| | | AC | 760 | 21,933 | 3.47% (3.22 – 3.71% | 905 | 28,258 | 3.20% (3.00 – 3.41%) |
| | | CC | 190 | 5,267 | 3.61% (3.10 – 4.11%) | 201 | 6,756 | 2.98% (2.57 – 3.38%) |
| | 70+ | AA | 1,125 | 38,842 | 2.90% (2.73 – 3.06%) | 936 | 44,617 | 2.10% (1.96 – 2.23%) |
| | | AC | 1,025 | 36,925 | 2.78% (2.61 – 2.94%) | 992 | 42,749 | 2.32% (2.18 – 2.46%) |
| | | CC | 267 | 8,694 | 3.07% (2.71 – 3.43%) | 262 | 10,210 | 2.57% (2.26 – 2.87%) |

Total subject numbers are restricted to subjects who were alive at the beginning of 2020. Age ranges refer to ages as of September 2020. Note that COVID rates by genotype in the overall

UKBB cohort are affected by subjects of non-White ethnicity tending to have higher COVID rates and lower C allele frequencies.
